# Investigation into the Release of Respiratory Aerosols by Brass Instruments and Mitigation Measures with Respect to Covid-19

**DOI:** 10.1101/2020.07.31.20165837

**Authors:** Alexander S. Parker, Kenneth Crookston

## Abstract

There are a number of recent studies detailing the transmission of SARS-CoV-2 (Covid-19) via both Droplet and Aerosol airborne particle routes of infection. Because of this, it is necessary to understand the release of different sized particles in activities such as playing brass instruments in order for an analysis of risk to take place for such activities.

In this investigation, the quantity and size of particles released by brass instruments while they are played was analysed for 7 different types of brass instrument. This was contrasted with the same individuals breathing as a comparison for more general activities as well as the effect of a mitigating polycotton barrier over the end of their instruments. To investigate the particles released, the particles were size sorted and counted with a six-channel laser particle counter. Multiple measurements were made by each individual in each condition investigated. The mean concentration exiting across all instruments measured was found to be 1.21×10^7^ ±1.03×10^6^ Aerosol type particles/m^3^ and 1.43×10^4^ ±9.01×10^2^ Droplet type particles/m^3^ per minute. When breathing, the mean count was 1.61×10^7^ ±1.33×10^6^ Aerosol type particle/m^3^ and 5.45×10^3^ ±1.20×10^3^ Droplet type particles/m^3^. When playing with a barrier cover, the mean number of particles emitted fell to 2.60×10^6^ ±2.11×10^5^ Aerosol type particle/m^3^ and 5.20×10^3^ ±8.02×10^2^ Droplet type particles/m^3^. This barrier represented an average 78.5% reduction for the number of respiratory Aerosol type particles and 63.8% reduction for Droplet type particles compared to playing an instrument without the barrier covering.

It was investigated what effect playing for a more extended period of time had on the release of particles with comparisons made to singing, breathing and covering the instruments’ bell ends with a barrier cap. This showed that the mean number of Aerosol type particles produced while playing was 5.38×10^7^ ±3.15×10^6^ Aerosol type particles/m^3^ produced and showed a significant drop in Aerosol type particle production when playing with a barrier used, with a mean average of 2.28×10^6^±8.01×10^4^ Aerosol type particles/m^3^. Both breathing and singing showed consistent numbers of Aerosol type particles produced with means of 6.59×10^7^ ±7.94×10^5^ Aerosol type particles/m^3^ and 5.28×10^7^ ±5.36×10^5^ Aerosol type particles/m^3^ respectively. This showed a drop in mean Aerosol type particles/m^3^ of 95.7% when using a barrier cap compared to playing without a barrier.

It is concluded that, while playing a brass instrument, the propagation of respiratory Aerosols does occur and, to a smaller extent, so do Droplet size particles, but at a lower level than when the subject was breathing without an instrument. Finally, it was shown that the use of a barrier cap on the bell end of the instrument offers a significant reduction in the production of respiratory Aerosols into the immediate surroundings, which offers a possible mitigation method for playing in groups from the release of Aerosol type particles, especially in hard to ventilate spaces.

**Funding:** This study was supported by funds from Arts Council England covering salary support for AP and KC. The cleanroom facility and particle counter were provisioned by Centre Stage Ltd. The funders did not have a role in the experimental design, data collection, analysis or decision to publish and content of the manuscript.

**Competing Interests:** The authors have declared working for Brass Bands England, which exists to support brass bands in England and the wider UK.

## Introduction

Due to the 2020 Pandemic of Novel Corona Virus SARS-CoV-2, understanding transmission of the virus in differing situations has become an area of acute interest[1,2]. There is evidence that certain events or activities have resulted in larger so called ‘superspreading’ events and understanding this transmission route is important in minimising risks[3–6].

In addition to contact transmission[7,8], when a virus is transmitted from contact with contaminated surfaces, one of the important mechanisms of transmission is via airborne particles[9,10] which, when brought into contact with membranous tissues (e.g. linings of the eyes, nose, mouth and lungs)[11–13] on susceptible individuals, allows the virus to enter and infect that individual[14,15]. These airborne particles are generally split into 2 categories, Droplets and Aerosols[16]. These airborne mechanisms are known to be important in the transmission of Tuberculosis and Measles[17,18]. Droplets are often defined as particles in the size of order larger than 5µm, which fall rapidly under the influence of gravity[19] and are thus fall within limited distances (≤1m). In contrast, Aerosols are defined as particles ≤5µm in size and remain suspended in the air for extended periods of time ranging from hours to days[20–23] and allowing them to spread over distances much greater than 1m[4]. One reason for the interest in airborne transmission is the presumed role of asymptomatic carriers in Covid-19 outbreaks[24]. There is also some evidence to suggest that airborne transmission is the dominant route for Covid-19[25].

Covid-19 has been shown to be transmitted as both Aerosol and Droplets[26], as well as in many other communicable diseases such as the Common Cold, Chickenpox, Rubella, Influenza, Pneumococcal Disease and Severe Acute Respiratory Syndrome (SARS)[18,27–31]. It has been shown this transmission is possible via normal breathing rather than requiring a cough or sneeze[32]. It should be noted, however, that this is less understood than contributions via contact methods of transmission[33]. There have, though, been documented cases of extremely contagious pathogens being transmitted via airborne particles, such as an outbreak of Measles in the USA that was traced to a sports gathering where, without evidence of close contact, transmission occurred between an athlete in the stadium centre to the public[34]. There is also a documented case of the spread of Tuberculosis within a ‘closed community’ military band, which concluded that the playing of instruments was a contributing factor to the spread of the pathogen[35]. This has implications for participants and audiences in other gatherings, as poor ventilation has been cited as a factor for increased risk of transmission[36–39].

Aerosols are generally formed when an infected individual coughs or sneezes but can also be made in more normal activity such as speaking or singing[39–43]. There has been research looking at the spread of viral particles using a Vuvuzela (a straight plastic blowing horn)[44] and microbial flora found in instruments[45,46], but in terms of musical instruments very little on Aerosol production has been peer-reviewed and published.

Due to the different sized airborne particles having different characteristics and that only a proportion of produced particles will contain pathogen material, analysis of the size of particles is crucial to understanding of how and activities ability to transmit diseases.

Brass bands have their origins in the early 19^th^ century in northern England[47]. The instruments are made from conical tubes of brass coiled for convenience of use, often around a set of valves used to add small lengths of tubing to alter the length. The length of tube can range depending on the exact instrument, in this work, starting with the cornet with ∼1.37m pipe length up to Bb Tuba with ∼5.5m. The instruments taper from a larger bell end (114-508 mm diameter) to a smaller mouthpiece (20-40mm diameter)[48]. We hypothesised that the combination of the length and coiling of the tubing might mitigate the propagation of particles that are collected in the condensate formed within the instrument. These instruments are traditionally used in the British-style brass band in community-based ensembles, playing at many celebration type events and gatherings within these communities[49]. Playing the instrument requires forcing air from the lungs between the players’ lips, forcing them to vibrate and set up a standing wave through the instrument[50]. As the instruments are commonly used in numbers of 25+ in situations involving gatherings of people, it is important to understand the extent to which airborne particle production takes place to assess the extent to which instruments might assist the spread of viruses like SARS-Cov-2. This can then be used to examine methods to minimise the spread of Covid-19 within playing ensembles, particularly indoors[51]. One measure commonly employed is the use of cloth masks which have been shown to be effective in limiting the release of Aerosols particularly in materials with higher weave density (thread count)[52,53].

## Results

### Instrumental comparison

To analyse the propagation of particles, 7 volunteers, each playing different brass instruments, were asked to play for 60 s periods of time to mimic the action of playing in a brass band as shown in Figure 1. To compare the effect of playing a brass instrument with no activity, subjects were also asked to breathe strongly into the particle counter collector funnel. To assess the possibility of a mitigating barrier a final test was carried out using a polycotton material cover over the instruments’ bell ends. Particles that exited the end of the instrument or mouth were counted using a Laser Particle Counter and classed as either Aerosol type particles, which were <5µm, or Droplet type particles >5µm, as these size differences are important in analysing the risk of viral transmission. Each scenario was carried out 5 times for the analysis and an average of these occurrences was used. Airborne particles were measured over a 60 s sample time while the subject was playing and recorded in particles/m^3^.

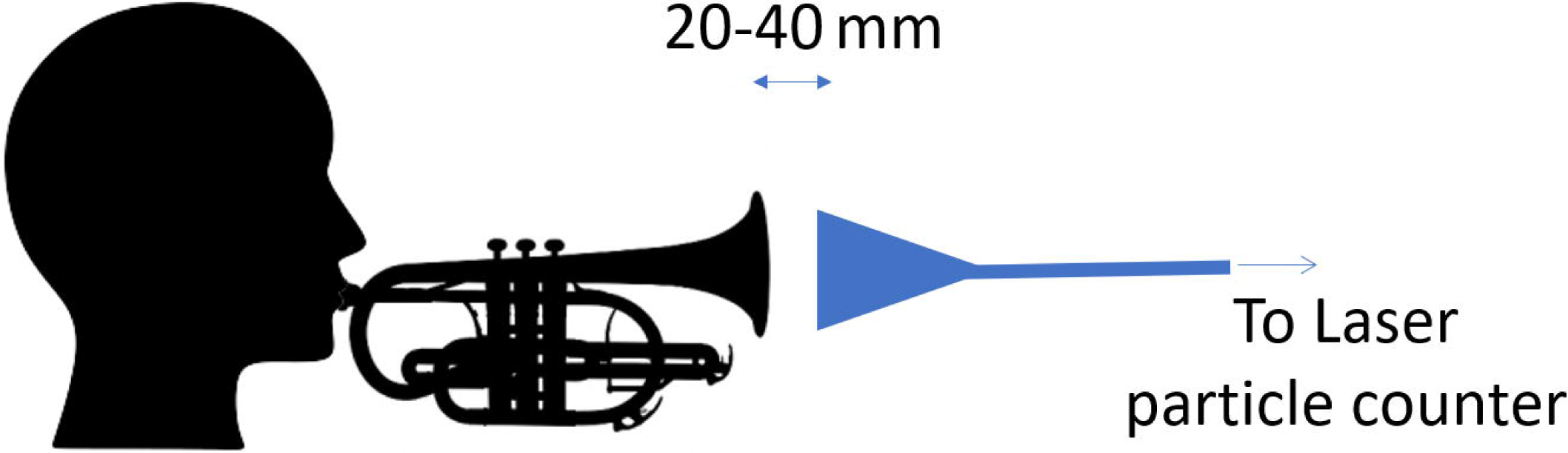

In each instrument test, both Aerosol and Droplet type particles were detected in measurable quantities above the background measurement. The mean average for each instrument can be seen in figure 2 for the release of Aerosol type particles and figure 3 for Droplet type particles. The mean concentration across all instruments measured was found to be 1.21×10^7^ ±1.03×10^6^ Aerosol type particles/m^3^ and 1.43×10^4^±9.01×10^2^ Droplet type particles/m^3^. The mean measurement for the same subjects when breathing was 1.61×10^7^±1.33×10^6^ Aerosol type particle/m^3^ and 5.45×10^3^±1.20×10^3^ Droplet type particles/m^3^. This shows that whilst both Droplets and Aerosols are produced while playing, they are produced differently. For Aerosols there are fewer particles produced while playing than whilst breathing, whereas for Droplets the reverse is true, but the difference is significantly smaller. When using the barrier there was a significant reduction in both types of particles with mean values of 2.60×10^6^±2.11×10^5^ Aerosol type particles/m^3^ and 5.20×10^3^±8.02×10^2^ Droplet type particles/m^3^. When compared to playing with no guard, this represents an Aerosol reduction of 78.5% and a Droplet reduction of 63.8%.

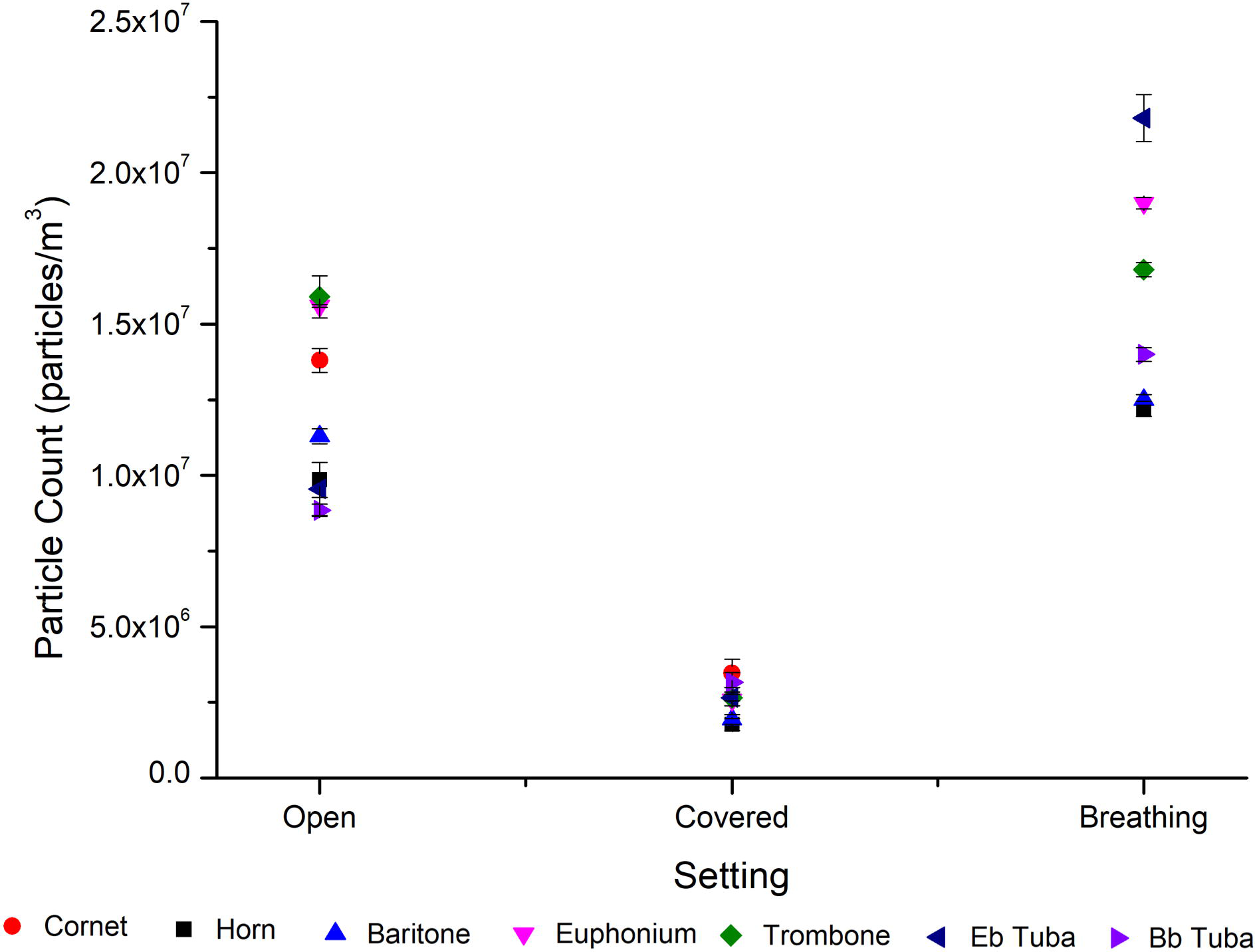

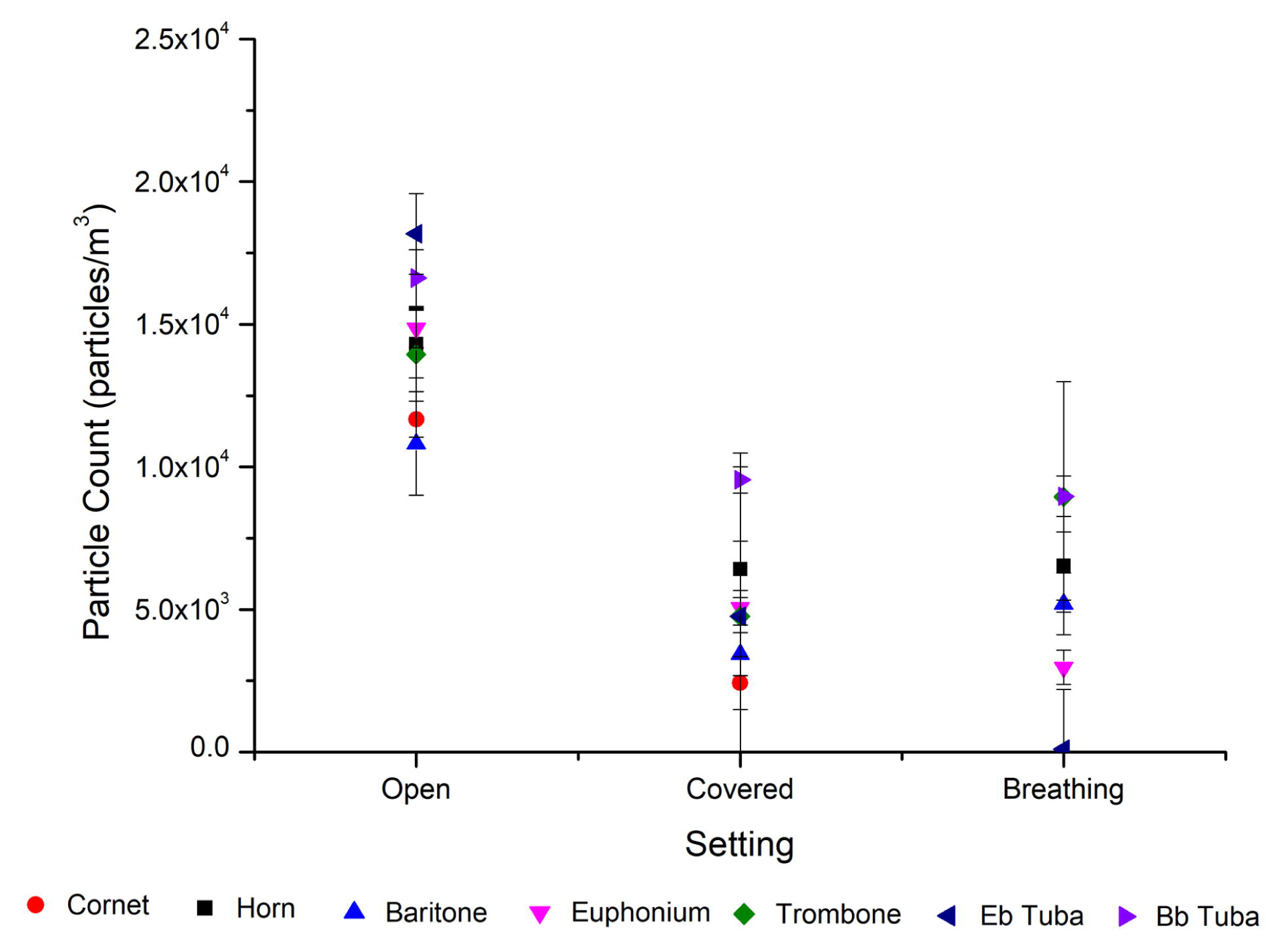

### Time resolved test

The subject was asked to play the instrument (Cornet) for the first 300 s at a medium volume, after which the test was repeated with the addition of a cloth barrier over the bell end of the instrument and again with the subject breathing into the collector and finally singing. The test was repeated 5 times with breaks for the subject to rest. When analysing the results, the particles were grouped into Aerosol type particles which were <5µm or Droplet type particles >5µm. The Droplet type particles were not found to be statistically significant and insignificant compared to the number of Aerosol type particles, which made up the vast majority of emitted particles, and were thus not used within the analysis. The result showing the mean average for the tests is shown in figure 4 and shows the mean average Aerosol type particles/m^3^ when the subject was playing their instrument at a medium volume, when playing at a medium volume with the barrier in place, when breathing strongly and when singing at a comfortable volume. Across all the samples when playing the instrument, the mean was 5.38×10^7^ ±3.15×10^6^ Aerosol type particles/m^3^ ; covered with the barrier mean was 2.28×10^6^ ±8.01×10^4^ ; breathing mean was 6.59×10^7^ ±7.94×10^5^ and singing mean was 5.28×10^7^ ±5.36×10 ^5^.

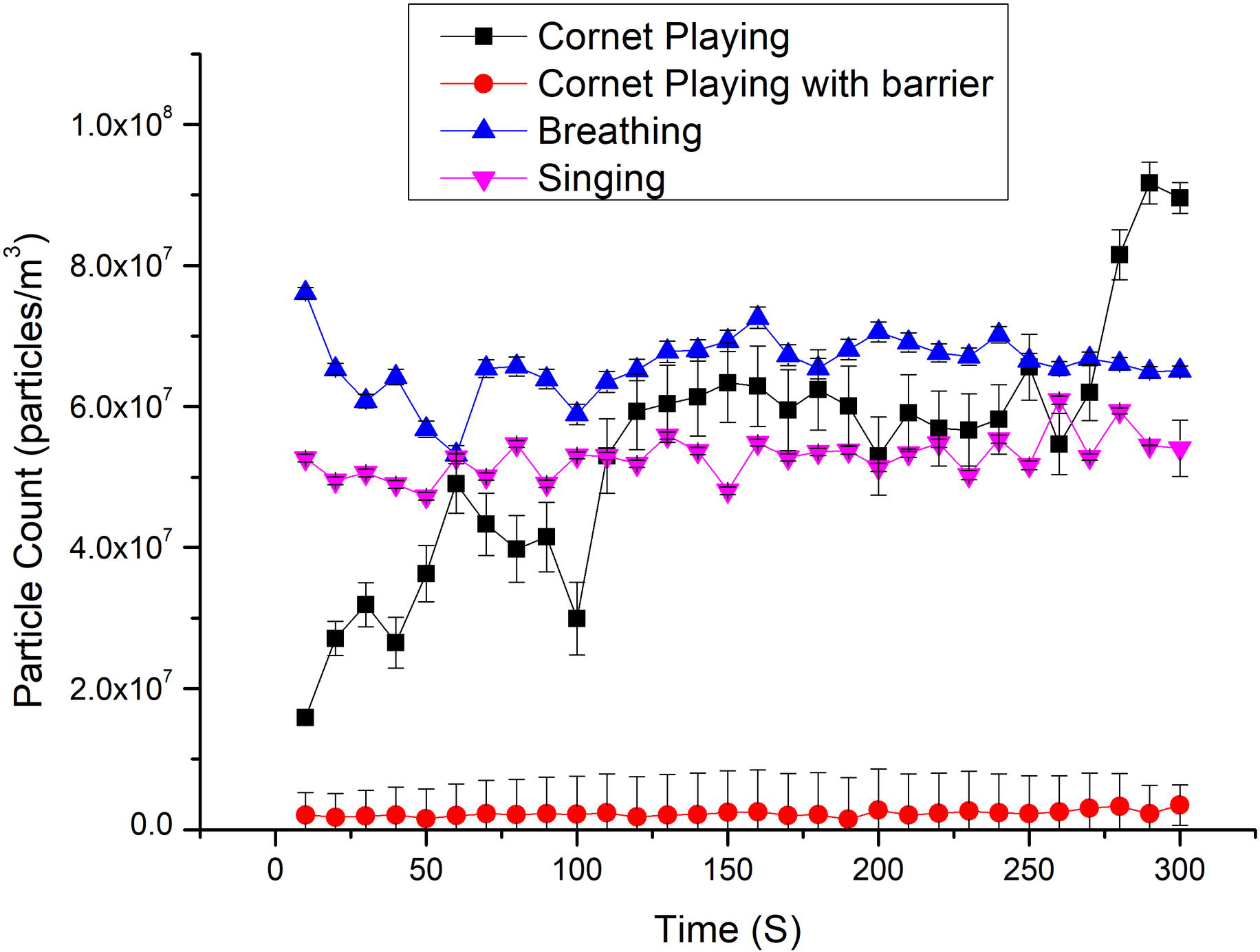

## Discussion

We have shown the number of both Aerosol and Droplet type particles propagated by 7 different brass instruments, showing that both particle types were produced, although significantly more Aerosol particles were emitted than the larger Droplet type particles. The mean concentration exiting the across all instruments measured was found to be 1.21×10^7^ Aerosol type particles/m^3^ and 1.43×10^4^ Droplet type particles/m^3^ per minute. The variation in particle release between different instruments appeared to be less significant than the difference between individual players as no discernible pattern related to either instrument size or design could be established. This was compared with mean measurement for the same subjects when breathing which was 1.61×10^7^ Aerosol type particle/m^3^ and 5.45×10^3^ Droplet type particles/m^3^. This shows that there were fewer particles emitted when playing than when the same subjects were breathing. There was a further and much more significant fall when playing with a barrier cover, with a mean number of particles emitted of 2.60×10^6^ Aerosol type particles/m^3^ and 5.20×10^3^ Droplet type particles/m^3^. This barrier represented an average 78.5% reduction for the number of respiratory Aerosol type particles and 63.8% reduction for Droplet type particles compared to playing and instrument without the barrier covering. It should also be noted that there was no statistical difference in the number of Droplet type particles emitted whilst playing with the barrier than when breathing. This result shows that whilst playing a brass instrument does propagate respiratory Aerosols, the number of particles is lower than heavy breathing. This may be because the instrument acts to partially filter the air flow, with Droplets collecting inside the instrument and expelled as condensate. It might also be that the coiling of the instrument and length of piping itself acts as a partial filter of particles, although further research would be needed to show this. With the use of a barrier, the number of particles was an order of magnitude lower showing a significant reduction in emitted Aerosols. This result suggests that due to the large number of Aerosol particles emitted, most of the particles are of a size that could remain suspended in the air as Droplet nuclei for extended periods of time if not properly screened with a barrier. These particles are of a size that could enter the alveoli space in the lungs[54].

With the time resolved test we showed that, over the course of the 300 s of playing, the number of particles showed an increasing trajectory rising from 1.59×10^7^ Aerosol type particles/m^3^ to a peak of 9.17×10^7^ Aerosol type particles/m^3^. The mean number of Aerosol type particles produced was 5.38×10^7^ Aerosol type particles/m^3^ produced. This would suggest that the particles take time to move through. This might be that 300 s of constant playing is not representative of normal playing where natural musical breaks allow for rest periods and the player expressed that the last part was difficult to maintain. This additional exertion might have resulted in additional Aerosol production within the final 30 s, which was a repeatable observation. It might also be a result of the fact that, while the subject could not see the instrument display while playing, they were told it was the final 30 s and they may have subconsciously made more effort. The result then showed a significant drop in Aerosol type particle production when playing with a barrier used with a mean average of 2.28×10^6^ Aerosol type particles/m^3^. This showed a drop in mean Aerosol type particles/m^3^ of 95.7%.

The number of particles produced does show a rise across the 300 s time of the experiment, but this was not significant when compared to playing without the barrier. Both breathing and singing showed consistent numbers of Aerosol type particles produced with means of 6.59×10^7^ Aerosol type particles/m^3^ and 5.28×10^7^ Aerosol type particles/m^3^ respectively. It was noted that fewer particles were produced for singing and playing than breathing, whilst playing with a barrier resulted in the significant fall in numbers by an order of magnitude. We hypothesise that the fall in particles for singing is a result of air flow control from the participant, lowering the air speed compared to breathing freely, although this would require further research to be confirmed. When playing a brass instrument, we also hypothesise the reduction in particles results from the coils and length of instrument tubing acts to capture emitted particles which forms into the condensate collected in the instrument. When playing with the barrier in place, it is clear that significant numbers of particles are captured in the barrier and are thus not emitted into the surroundings. Particles that are emitted would, due to their size, remain suspended for significant amounts of time and if inhaled would be capable of penetrating the respiratory system to the alveoli level[54]. This would indicate that the use of this barrier is effective in reducing the particle release while playing and should be considered in any assessment of the risks associated with playing in groups. This is particularly true in poorly ventilated or indoor spaces where particles would remain for extended periods of time and reducing emission would be particularly valuable.

Within this test there was not scope to test other potential areas of particle emission, such as the junction between the players’ lips and the mouthpiece, although there was no observable release of air when properly played. Another potential area is the emptying of collected condensate. Although this was an infrequent practice in these experiments, this would require further investigation. This investigation was also not able to measure the volume of air players inhaled and exhaled whilst playing as this will also have an effect of the risk of viral transmission linked to the number of potential particles inhaled[12,13]. However, reducing the number of particles in the local atmosphere will play a part on reducing this risk. It should also be noted that, due to ethical reasons, only healthy volunteers were used for this investigation, but that particle size and number can be affected depending on the pathogen a subject might have[55–57]. A patient with impaired lung function as a result of a pathogen might also produce different result due to reduced ability to move air effectively.

It has been shown that while playing a brass instrument the propagation of respiratory Aerosols does occur and to a smaller extent so do Droplet size particles. It is shown that when the same subject is breathing under otherwise identical conditions, a higher number of particles are produced than when playing. It has also been shown that the use of a barrier cap on the bell end of the instrument offers a significant reduction in the production of respiratory Aerosols into the immediate surroundings. This would indicate that the barrier cap should be included within any assessment of risk while playing as a mitigation measure against the release of airborne particles.

## Materials and Methods

This study was carried out at Modular Sterile Developments (Unit 1B, Dale Mill, Burnley Road East, Rossendale, BB4 9HU Tel: 01706 231598) using a clean room with ISO Class number 7. Ethical approval was obtained from Brass Bands England’s Board and informed consent was obtained in writing from all participants. Eight healthy volunteers covering 7 different brass instruments participated in the research. The test subjects used in the tests were in the age range 30-50 and self-reported as free from illness. The instruments played as part of the experiments were Cornet, Tenor Horn, Baritone Horn, Euphonium, Trombone, Eb Tuba and Bb Tuba. All players and singers were of a professional standard. To avoid cross-contamination, each participant used their own instrument. As part of the test, a cloth cover was tested as a barrier to emitted particles. These were provided by Centre Stage Uniforms Ltd. and consisted of an elasticated cap to attach tightly over the end of the instrument bell and made from a woven polycotton material (50/50 poly cotton, 76/68, 30/30). Particles were measured using a Lasair III 310C six-channel laser particle counter (EMS Particle Solutions UK Ltd). During the experiment, particles were collected and counted in 6 categories according to their diameter 0.3-0.5µm; 0.5-1µm;1-5µm; 5-10µm; 10-25µm and >25µm.

### Instrument comparisons

For the measurements, the subjects were left alone in the clean room after the technician left the room. After a 60s period for the room to settle, the instrument would begin sampling. Subjects were asked to play continuously apart from breathing at a natural rate and to play at a medium comfortable dynamic as close to how they would play normally in a band setting for a sampling time of 60 s. It was considered that 60 s was a representative time for playing in an ensemble setting. For each sample 0.0283 m^3^ air was collected and the particles counted. The subjects were then given a 20-second break before repeating the experiment. For each participant subject there was a background reading taken followed by playing with the bell barrier, playing without the barrier and with the subject breathing with no instrument. For each scenario, 5 repeat measurements were made. The subjects were not able to see the instrument displays while the samples were being taken and were instructed when to play and rest via instruction through a window into the clean room from the technician. The player held the bell end of the instrument (or mouth when breathing) approximately 20-40 mm from the collection funnel of the particle counter inlet funnel.

### Time resolved experiment

For the time resolved experiment, a single participant was used playing a cornet. In this test the subject was also alone in the clean room and, after a 60 s time period for the room to settle, began to play. The subject was instructed to play in the middle register at a medium dynamic and aiming to play 2 notes per second. Samples were taken every 10 s over a period of 300 s. For each sample 0.0047 m^3^ of air was collected and the particles counted. This was repeated 5 times in each scenario of playing with the barrier, playing without the barrier, singing and breathing. The player held the bell end of the instrument (or placed mouth when breathing or singing) approximately 20-40 mm from the collection funnel of the particle counter inlet. It was noted that 300 s was an extreme length of time for constant playing as musical breaks usually allow for short rest periods.

Analysis was made on the total sample particle count for each particle size and sample volume adjusted to a standard particle count/m^3^ for comparison. For the instrument comparison experiments, particles were classed as either Aerosol type particles (<5µm) or Droplet type particles (>5µm) as these particle sizes are of most interest with regards to differences in potential viral transmission routes. For all experiments, the samples were corrected to remove the background measurement for each participant in order to give a like-for-like comparison of particles emitted. For the time resolved experiments, the Aerosol type particles only were used in the analysis as the larger Droplet type particles were found to make up a negligible part of the total count.

## Data Availability

All relevant data are within the manuscript and its Supporting Information files.

## Acknowledgments

The Authors would like to thank Centre Stage Ltd. for the provision of the barrier covers. We are grateful to the volunteers who supported this study.

## Author Contributions

The experimental conception and design: AP. Performed the Experiments: AP. Analysed data: AP. Wrote the paper: AP KC.

